# autoscoRA: Deep Learning to Automate Sharp/van der Heijde Scoring of Radiographic Damage in Rheumatoid Arthritis

**DOI:** 10.64898/2025.12.26.25343056

**Authors:** Thomas Deimel, Paul J. Weiser, Martin Urschler, Christian Payer, Peter Mandl, Georg Langs, Daniel Aletaha

**Author notes:** Authors contributed equally (Thomas Deimel, Paul J. Weiser).

## Abstract

**Objective:** Regular imaging by conventional radiography to assess for joint damage is a cornerstone in the management of rheumatoid arthritis (RA). Scoring systems to quantify such damage, such as the widely used Sharp/van der Heijde (SvdH) score, are limited by the requirement of time and experienced staff as well as intra- and inter-rater variability. To alleviate these problems, autoscoRA, a fully automated scoring system to assign SvdH scores to radiographs of the hands and feet was developed.

**Methods:** Using the hitherto largest dataset of adult rheumatoid arthritis patients, autoscoRA, a deep learning-based system, was trained to automatically perform joint extraction and scoring of joint space narrowing and bone erosion.

**Results:** The dataset included 769 patients (155 of which in the test set) with 3437 visits (707) and 12144 radiographs (2507). The model reached excellent agreement with a human scorer for joint space narrowing, erosion, and combined scores both on the joint level and for summed total SvdH scores (ICC 0.9). On a subset of data scored by a second human reader, the model outperformed the former in terms of agreement with the first human reader. In addition, autoscoRA demonstrated good agreement with a human reader for detecting longitudinal progression of joint damage across different SvdH score cut-offs defining the presence of progression (average agreement of 70 %).

**Conclusion:** Automated systems like autoscoRA could be used to facilitate scoring of radiographic joint damage in clinical trials, registries and observational studies, and, eventually, routine clinical care.

## Introduction

Rheumatoid arthritis (RA) is a systemic immune-mediated inflammatory disease, which most commonly presents as a symmetric, peripheral polyarthritis^1^. Chronic inflammation of the synovial membrane lining the inner surface of joint capsules is generally viewed as a leading cause of irreversible destruction of bone and degradation of cartilage^2^. Therefore, management strategies of RA focus on abating inflammatory disease activity to preserve structural integrity and functional capacity in individuals affected by RA. Whether this strategy is successful can be confirmed by periodical (typically annual) imaging of the hands and feet by conventional radiography^3,4^. Several comprehensive and sensitive tools, such as the widely used Sharp/van der Heijde (SvdH) score^5^, have been developed to quantify the acquired structural joint damage^6^. However, application of these scores is time-consuming and requires specially trained and experienced staff, and results are therefore also subject to significant intra- and inter-rater variability^6,7^. Together, these limitations pose problems for the use of semi-quantitative scores in both research and daily rheumatological practice. In fact, semi-quantitative scoring systems have shown a lack of feasibility in routine clinical care, essentially limiting their use to clinical trials.

Potential solutions are automated scoring systems, which could alleviate both practical and reliability-related issues. Most “manual” scoring systems in use today assign scores to a large number of joints according to at least two criteria: joint space narrowing (JSN) (representing cartilage damage) as well as the presence and degree of bone erosions^6^. Early attempts at automation have largely focused on (semi-)automatic measurement of joint space widths^8–13^. In addition, a smaller number of developers have created systems designed to (additionally) detect and grade bone erosions^14–16^. None of these early systems, however, have been able to reach a level of accuracy, reliability, and practicality to allow large-scale replacement of manual scores. Furthermore, most of these attempts relied on very small data sets and were limited by the more traditional computational methods used at the time.

Developments in the areas of computer vision and machine learning—most notably the introduction of so-called deep convolutional neural networks^17,18^, which have been used in various fields including medical imaging^19–22^—have brought an automated scoring of radiographic damage within reach. Recent experiments have demonstrated increasingly promising results^23^. However, these approaches are constrained by the use of small datasets, the exclusion of foot joints, the absence of automated patch extraction techniques, or a focus solely on predicting JSN or erosion^24–31^.

Radiographic joint damage remains a cornerstone outcome in RA, both for guiding long-term patient management and as a pivotal endpoint in clinical trials, while its gold-standard assessment by the Sharp/van der Heijde (SvdH) score is limited by time demands, the need for expert readers, and considerable inter- and intra-observer variability. An automated system such as autoscoRA directly addresses the feasibility gap, offering a scalable and reproducible solution that transforms imaging into reliable, structured outcome data.

## Methods

The project was approved by the Ethics Committee of the Medical University of Vienna (EC No. 1206/2018). The model implementation is openly available on GitHub^a^, a user interface to allow clinicians to directly apply the model is currently not available.

### Patients and data

All consecutive adult RA patients managed at a large academic medical center between the years 2000 and 2018 were included in this study if they had given prior consent to use of their clinical data and if appropriate radiographs were available in the hospital’s picture archiving and communication system (PACS). Patients with additional other inflammatory joint diseases were excluded. Patients had undergone regular radiographic monitoring of disease progression as part of routine care. A given point in time where radiographs were taken is referred to as a visit. A visit with a complete set of radiographs is a visit for which dorsopalmar/-plantar radiographs of both hands and both feet (i.e., all images needed to calculate the total SvdH score) are available. Radiographs were extracted from the hospital’s PACS and pseudonymized. Inadequate images (e.g., not depicting feet or hands or not showing all joints contained in the SvdH score) were discarded, issues like malrotation were corrected, and right extremities were mirrored.

The complete dataset was split into training (60 % of patients), validation (20 %), and test set (20 %) on the patient level (i.e., all radiographs of a given patient were contained in only one of the three sets). The training set was used to fit model parameters and the validation set for choosing the best-performing model (i.e., the optimal model architecture and hyperparameter settings). The chosen final model’s performance was then evaluated on the independent test set. Descriptive statistics on the patient and visit level were calculated. Differences between the three datasets were tested for using ANOVA and Kruskall-Wallis tests for means and medians, respectively.

SvdH scores were assigned by an expert reader with more than 10 years of experience in scoring radiographs according to the SvdH method^5^. The reader was allowed to view consecutive radiographs side by side and was not blinded to time order. For a subset of images, SvdH scores from a second reader, who had received training from the first reader over several months, were available from an unrelated previous project.

The SvdH system uses dorsopalmar/-plantar radiographs of both hands and both feet. It assigns a separate score for erosions (0-5 for hand joints, 0-10 for foot joints) and JSN (0-4, where 0 = normal, 1 = focal or doubtful, 2 = generalized but > 50 % of joint space left, 3 = subluxation or < 50 % of original joint space left, 4 = luxation or bony ankylosis), respectively, to each assessed joint. These scores are then summed up to a total score (0-448). The erosion score indicates the degree of bony damage, with 1 being a discrete erosion and 5 indicating complete collapse of the bone. For finger and toe joints, the erosion score is first assigned separately to each of the two joint surfaces (each ranging from 0 to 5) and then added together. For the finger joints, the summed erosion score for each joint is capped at 5, while the toe joints may have erosions scores up to 10^5^.

### Model development

Model development was performed in a cross-sectional manner, with a radiograph (or rather, region of interest around a specific joint) from a given visit being used as input and the predicted SvdH joint score as output.

#### Joint localization

Automatic joint localization was performed using a deep learning approach dubbed SpatialConfiguration-Net, initially developed on datasets of pediatric hand radiographs and magnetic resonance images^32^. In brief, this convolutional neural network combines information about the local appearance of a specific joint with its expected location relative to the predicted localization of other joints in the image. The model was trained separately on 100 hand and 100 foot images (belonging to the training set) and corresponding manually annotated joint locations. Localization accuracy was assessed using three-fold cross-validation, filtering for images with no operated joints and with available pixel spacing values, i.e., the resolution in millimeters per pixel. The trained model was then applied to all remaining images of the training set, as well as to the validation and test sets.

For metatarsophalangeal (MTP), metacarpophalangeal (MCP), proximal interphalangeal (PIP), and first digit interphalangeal (IP) joints, the tip of the proximal bone and the base of the distal bone forming the joint were localized separately, as separate SvdH erosion subscores were available for the proximal and distal components of these joints, respectively. For the carpal bones/joints contained in the SvdH score, only the center of the wrist was localized. In addition to the actual joints of interest, helper points were localized that would aid in determining the axis, size, and position of regions of interest (see below) around joints.

#### ROI extraction

Regions of interest (ROIs) around the joints located in the previous step were extracted automatically. These regions of interest were later used as the input to the deep learning model assigning automated scores to each joint. For all except the wrist and CMC joints, two ROIs were extracted per joint, one for the proximal and one for the distal joint surface. For CMCs, one ROI was extracted per joint. For the wrist, only one ROI was extracted containing all wrist joints/bones, which was later used as input in the training of each wrist joint/bone’s individual model, respectively. Square-shaped regions of interest, as used here, are defined by their location, orientation (axis), and size (side length). ROI orientation was defined by the axis of the corresponding bone, as obtained from automatically localized points at the proximal and distal bone tips. To account for inter-individual differences in hand/foot sizes as well as the fact that the resolution in millimeters per pixel was not known for all radiographs, ROI sizes were scaled based on a “reference size” λ, designed to represent the overall size of an extremity. This reference size was obtained by taking the median of the lengths of the metacarpal, proximal phalangeal, and intermediate phalangeal bones II-V (for hands), and metatarsal and proximal phalangeal bones I-V as well as the distal phalangeal bone I (for feet), respectively. Bone lengths, similarly to ROI orientation, were determined by utilizing automatically localized proximal and distal bone tips. Since joint types differ in size, a linear model (with intercept set to zero) relating λ to ROI size was fit for each joint type. Similarly, models predicting the shift of an ROI center from a previously determined joint position were fit for each joint type. ROI extraction models were fit on the same 100 hand and 100 foot images previously used for training automated joint localization. The trained models were then applied to all remaining images of the training set, as well as to the validation and test sets.

[VALIDATION PART OF THIS SUBSECTION FUSED INTO MODEL VALIDATION SECTION OF METHODS]

#### Training of the scoring model on the training set

To maximize the available training data and increase generalizability, data augmentation was performed by applying small random transformations to ROIs both before (translation, rotation, and rescaling of the ROI outline) and after (translation, rotation, rescaling, shearing, elastic deformation, and addition of random gaussian noise using MONAI^33^ on the extracted ROI image) the actual extraction of ROIs. Upsampling was applied to balance the number of samples in each class.

Separate scoring models were trained for each joint group (CMCs, MCPs, PIPs including IP of the hand, MTPs, IP of the foot) and each individual bone/joint of the wrist. For non-wrist erosion prediction, separate models were trained for the proximal and distal joint surfaces, the predictions of which were subsequently added up to obtain the joint erosion score (with an upper limit of 5 for finger and an upper limit of 10 for toe joints as in the SvdH score). Target label were the joint-level scores (or joint surface-level scores for non-wrist erosion scores) assigned by the human reader and will be referred to as “Ground truth labels”.

All models were based on the VGG-19 architecture^34^. We compared training as regression (mean squared error loss function), ordinal regression^35^, classification (cross entropy loss function), and penalized classification. For the latter, a cross entropy-based loss function was extended by a term penalizing larger deviations from the ground truth score to yield

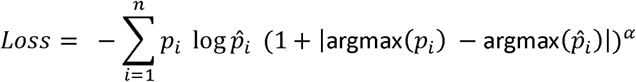

where *p_i_* is equal to 1 when class *i* corresponds to the ground truth label and 0 otherwise, 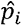 represents the predicted probability of class *i*, and *a* is an adjustable parameter regulating the strength of penalty applied. Values for *a* were tested within a range of 0 to 1 with a step size of 0.2. All models were initially trained for 50 epochs on the training set using the Adam optimizer^36^ with a learning rate of 0.0005. The best model weights were chosen based on the validation set loss. For classification, the loss on the validation set was computed by taking the mean across classes.

### Model validation

With the exception of testing the model’s ability to assess radiographic progression, model validation was performed in a cross-sectional manner, with an ROI from a given visit being used as input and the predicted SvdH joint score as output.

#### Joint localization and ROI extraction

The performance evaluation of automated localization and ROI extraction was conducted by three-fold cross-validation, filtering for images that showed no operated joints (and, for the localization task, had available pixel spacing values, i.e., the resolution in millimeters per pixel). Performance of the localization tasked was assessed using the deviation in mm from manually annotated joint locations. For ROI extraction, all automatically extracted ROIs were manually evaluated based on whether the entirety of the joint space or bone surface of interest was depicted (no “crop”), on whether parts of any adjacent joint spaces were visible (“overlap”), and for malrotation of more than 30°. The evaluation was performed on each group of joints including the wrist, CMC, MCP PIP, MTP and IP joints. ROI extraction was evaluated based on manually annotated joint locations to evaluate its performance independently from the localization task.

#### Scoring model selection

The final model was manually chosen based on the performance on the validation set, using several statistics including accuracy, accuracy if any deviation of less than 2 was considered correct, balanced accuracy, balanced accuracy if any deviation of less than 2 was considered correct, and two-way mixed intraclass correlation ICC(3, 1) with single measurement on the joint level, as well as ICC(3, 1) for scores summed within and across extremities and/or score domains (i.e., JSN or erosion). In addition, visual inspection of the joint-level results for robustness and plausibility of predictions across joints was performed.

#### Scoring model validation

Performance of the chosen final model was assessed in detail by applying it to the independent test data set. For joint level score predictions, confusion matrices were drawn, and accuracy, balanced accuracy, the percentage of joints with a deviation from the ground truth score of more than 1, as well as balanced accuracy if any deviation of less than 2 was considered correct were calculated. For the total SvdH score as well as for the total JSN and erosion scores, scatter plots were drawn, and spearmen correlation and ICC(3, 1) were computed. The same graphical representations were created and statistical measures were calculated when comparing the automated scoring system to a second human reader on a subset of the test dataset.

#### Detection of radiographic progression

Changes in the automatically assigned score over time were calculated by subtracting a score at an earlier visit from that at a later visit of the same patient. These score differences were compared to those between the corresponding scores assigned by the reference human reader. All possible (including non-consecutive) pairs of visits with complete sets of radiographs that were conducted at least five years apart for each patient within the test data set were used. The five-year gap was set to allow for a fairer comparison between the model and the reference human reader. Similarly to radiologists, the human reader assigned scores by looking at two consecutive radiographs simultaneously (“parallel scoring”), while the automated scoring system only sees radiographs from one particular visit at a time. Thus, the latter is at a disadvantage in detecting radiographic progression.

Performance was assessed over a range of definitions of radiographic progressions (reference human reader-assigned increase in the SvdH score between 0 and 20 in steps of 1). The number of false positive, true negative, true positive, and false negative cases as well as specificity and sensitivity were recorded for each cutoff.

## Results

### Patients and data

In total, 769 patients (155 of which in the test set) with 3437 visits (707) and 12144 radiographs (2507) were available for model development and evaluation. Error! Reference source not found. displays the characteristics of patients in the training, validation, and test datasets. The distribution of SvdH scores for each of the three datasets is shown in Supplementary Figure 1.

### An automated SvdH scoring system

The three-step setup of our system is depicted schematically in Error! Reference source not found.. As the final scoring model to be applied to the test dataset, a VGG-19 convolution neural network trained with a penalized cross-entropy loss function with an of 0.8 was chosen. The performance of the automated scoring system on the unseen test set is presented in Error! Reference source not found.. Panel B shows confusion matrices displaying the agreement between human reader- and automatically generated scores on the joint level, where an accumulation of scores along the diagonal indicates good agreement. There was an excellent performance on JSN for joints of both the hands and feet, with misclassifications that deviated by more than a score difference of 1 from the human reader ground truth occurring in only 2.6 and 3.0 % of cases for hands and feet, respectively. For erosions, agreement was good as well, but scores appeared somewhat more spread around the diagonal. Of course, the definition of the erosion score in the original SvdH scoring system also allows for a wider range of scores on the joint level than for the JSN part of score. In particular, for the feet, the joint-level erosion score can take values between 0 and 10, while the JSN score only allows for values between 0 and 4. Misclassifications that deviated by more than 1 from the human reader ground truth erosion score still occurred in only a small proportion of 6.3 and 11.0 % of cases for hands and feet, respectively. Detailed performance measures are depicted in Table 2. Performance across different levels of disease severity is displayed in Supplementary Table 1.

**Table 1.** Overview of patient characteristics for training, validation, and test set. ANOVA (or Kruskal-Wallis) tests were used to test for differences in the means (or medians) of a variable between the three datasets. Complete set of radiographs, visit for which radiographs of all four extremities are available, i.e., left and right hands and feet; sd, standard deviation.

|  | overall | training | validation | test | p value | statistical test |
| --- | --- | --- | --- | --- | --- | --- |
| **patients** | **769** | **463** | **151** | **155** |  |  |
| patients with $\geq 1$ visit with a complete set of radiographs* (%) | 643 (83.6) | 382 (82.5) | 128 (84.8) | 133 (85.8) | | |
| mean (sd) age at last visit (years) | 60.5 (13.3) | 60.4 (13.2) | 61.2 (13.4) | 60.3 (13.5) | 0.28 | anova |
| female sex (%) | 598 (77.8) | 359 (77.5) | 119 (78.8) | 120 (77.4) | 0.94 | chi squared |
| seropositive (%) | 522 (67.9) | 319 (68.9) | 100 (66.2) | 103 (66.5) | 0.76 | chi squared |
| mean (sd) disease duration at last visit (years) | 11.6 (9.9) | 12 (10.3) | 11 (9.2) | 10.8 (8.9) | 0.28 | anova |
| mean (sd) SvdH score at last visit | 35.5 (44) | 37.2 (45.6) | 37.3 (46.1) | 29.1 (36.1) | 0.16 | anova |
| mean (sd) erosion score at last visit | 11.1 (19.9) | 11.9 (20.5) | 11.5 (21.7) | 8.4 (15.8) | 0.22 | anova |
| mean (sd) JSN score at last visit | 24.4 (26.2) | 25.3 (27.1) | 25.8 (26.8) | 20.7 (22.5) | 0.17 | anova |
| mean (sd) estimated annual progression | 2.6 (3.2) | 2.7 (3.5) | 2.6 (2.8) | 2.1 (2.7) | 0.31 | anova |
| **visits** | **3437** | **2056** | **674** | **707** |  |  |
| visits with a complete set of radiographs* | 2453 (71.4) | 1461 (71.1) | 478 (70.9) | 514 (72.7) |  |  |
| median (min, max) visits per patient | 4 (1, 14) | 4 (1, 14) | 4 (1, 13) | 4 (1, 14) | 0.68 | kruskal-wallis |
| median (min, max) visits with a complete set of radiographs* per patient | 3 (0, 12) | 3 (0, 12) | 3 (0, 10) | 3 (0, 10) | 0.65 | kruskal-wallis |
| median (min, max) years of followup | 4.6 (0, 16.8) | 4.5 (0, 16.8) | 3.9 (0, 15.4) | 5.1 (0, 16) | 0.23 | kruskal-wallis |
| **radiographs** | **12144** | **7282** | **2355** | **2507** |  |  |
| hand radiographs | 6318 | 3759 | 1232 | 1327 |  |  |
| foot radiographs | 5826 | 3523 | 1123 | 1180 |  |  |
| **radiographs by x-ray vendor** |  |  |  |  |  |  |
| Philips | 7776 | 4591 | 1489 | 1696 |  |  |
| Fuji | 2548 | 1611 | 467 | 470 |  |  |
| AGFA | 919 | 560 | 183 | 176 |  |  |
| Siemens | 359 | 196 | 90 | 73 |  |  |
| VIDAR = scanned | 542 | 324 | 126 | 92 |  |  |

**Table 2.** Performance statistics of the automated system compared to ground truth human reader at the joint level. Error > 1, percentage of joint-level scores with a deviation of > 1 from the ground truth score; balanced acc. (error < 2), balanced accuracy if a deviation of 1 from the ground truth score was accepted; RMSE, root mean square error.

|  | joints | accuracy (%) | error > 1 (%) | balanced acc. (%) | balanced acc. (error < 2) (%) | RMSE |
| --- | --- | --- | --- | --- | --- | --- |
| erosion hand | 21232 | 79.4 | 6.3 | 41.1 | 81.7 | 0.72 |
| erosion foot | 7080 | 75.3 | 11.0 | 23.9 | 58.2 | 0.96 |
| JSN hand | 19905 | 72.4 | 2.6 | 61.3 | 96.4 | 0.61 |
| JSN foot | 7080 | 75.6 | 3.0 | 61.7 | 95.2 | 0.60 |

Note that differences in overall prevalences of scores between the human reader and automated scoring system (see the bar plots on top and to the right of each confusion matrix, respectively) are, at least in part, inherent to the imbalanced number represented at each score level. Indeed, inspecting joint level predictions for a given ground truth score, predictions seem to fall approximately symmetrically around the ‘true’ value. However, due to the imbalance of the score distribution, overestimation of lower ground truth scores contributed higher absolute numbers to the distribution of predicted scores than an underestimation of higher ground truth scores. Thus, if predictions are distributed symmetrically at each score level, their overall distribution is necessarily shifted toward higher scores.

In panel A of Error! Reference source not found., the association of total SvdH, total JSN, and total erosion scores between the human reader and the automated scoring system is displayed. Overall, there was a strong association as indicated by high values for Spearman correlation and intraclass correlation (ICC). There appears to be a certain bias towards higher values for both the total erosion and, in consequence, the total SvdH score. This is, at least partially, a direct consequence of the differences in score prevalence between the human reader and the automated scoring described above and potentiated during the summation process.

### Human vs. machine learning

Supplementary Figures 2 and 3 display a comparison of human and machine performance. A subset of 68 foot and 72 hand radiographs of 35 and 38 patients, respectively, including 31 complete radiographic sets (two hands and two feet) of 31 patients were scored by a second reader. Agreement with the first reader (ground truth) was assessed for both the automated scoring system and the second human reader. Supplementary Figures 2 and 3 demonstrate the superior performance of the automated scoring system compared to the second human scorer for summed scores and individual joint scores, respectively. For summed scores, the automated scoring system achieved an ICC of 0.94 compared to the second reader’s ICC of 0.86. On the joint level, misclassifications that deviated by more than a score difference of 1 from the first human reader ground truth JSN score occurred in only 2.2 and 2.6 % of cases for the automated scoring of hand and foot joints, respectively. For the second human reader, these numbers were 10.9 and 9.3 %, respectively. For the erosion score, the performance of the automated system roughly matched that of the second human reader numerically, although visual inspection indicated potentially more consistent predictions by the former. Detailed performance measures are depicted in Table 3.

**Table 3.** Performance statistics of the automated system and second human reader compared to ground truth human reader at the joint level. Error > 1, percentage of joint-level scores with a deviation of > 1 from the ground truth score; balanced acc. (error < 2), balanced accuracy if a deviation of 1 from the ground truth score was accepted; RMSE, root mean square error.

|  |  | joints | accuracy (%) | error > 1 (%) | balanced acc. (%) | balanced acc. (error < 2) (%) | RMSE |
| --- | --- | --- | --- | --- | --- | --- | --- |
| second reader | erosion hand | 1152 | 0.87 | 0.04 | 0.26 | 0.62 | 0.61 |
| second reader | erosion foot | 408 | 0.78 | 0.09 | 0.17 | 0.55 | 0.94 |
| second reader | JSN hand | 1080 | 0.58 | 0.11 | 0.60 | 0.95 | 0.88 |
| second reader | JSN foot | 408 | 0.65 | 0.09 | 0.43 | 0.87 | 0.81 |
| autoscoRA | erosion hand | 1152 | 0.82 | 0.05 | 0.29 | 0.74 | 0.66 |
| autoscoRA | erosion foot | 408 | 0.78 | 0.09 | 0.23 | 0.55 | 0.97 |
| autoscoRA | JSN hand | 1080 | 0.74 | 0.02 | 0.66 | 0.97 | 0.58 |
| autoscoRA | JSN foot | 408 | 0.79 | 0.03 | 0.57 | 0.93 | 0.56 |

### Detecting radiographic progression

Results for the model’s ability to detect radiographic progression are displayed in Figure 3 and include 54 patients with a total of 237 visits. The results show an average agreement of 70 % across the different progression-defining increases in SvdH scores. Overall performance appeared to be relatively stable across the range of cutoffs. Performance in subsets with low and high baseline joint damage, respectively, can be found in Supplementary Figure 4.

**Figure 1.**
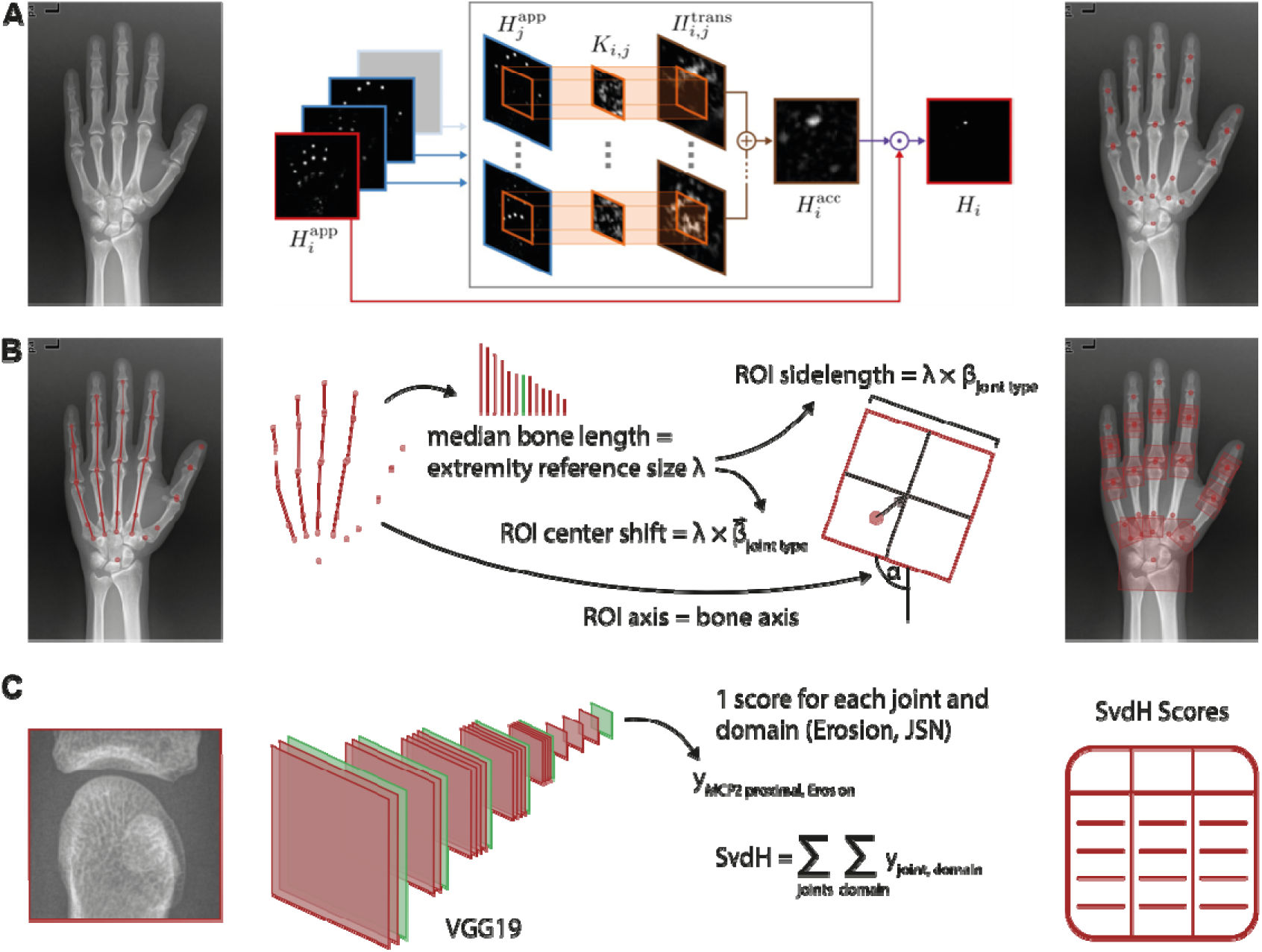
Schematic display of the three-step approach to automate SvdH scoring of radiographic damage. A. Joint localization. Joints in plain radiographs of hands and feet are automatically localized using a SpatialConfiguration-Net (SCN)^32^ that combines information of local appearance of a given joint with such on the spatial relationship to all other joints. B. Region of interest (ROI) extraction. The median of bone lengths obtained from joint localizations is used as a reference size λ for each extremity. This quantity is then used to determine the side length of each square-shaped ROI as well as the shift of the ROI center from the previously determined joint positions by fitting linear models with intercepts set to zero. ROI orientation was set to the orientation of the adjacent bone as obtained from joint localizations. C. SvdH scoring. Regions of interest are extracted and used as the input for a VGG19 convolutional neural network trained on human reader-generated joint-level scores for erosion and JSN. Joint-level scores are summed up according to the rules of the SvdH scoring system to obtain total scores for a given timepoint and patient.

**Figure 2.**
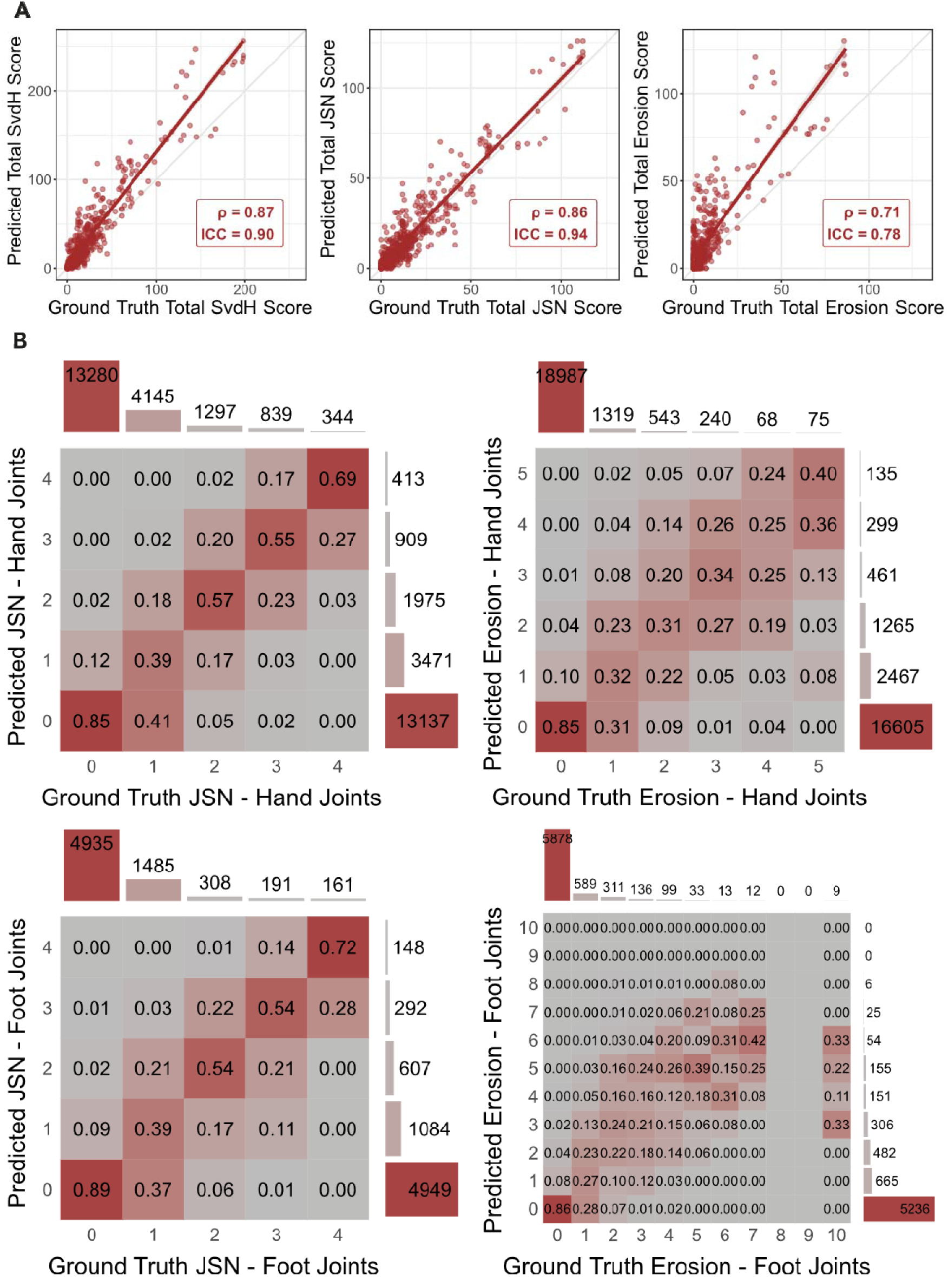
Test set comparison of the automated scoring with ground truth manual SvdH scores. A. Scatter plot of sum scores, including the total SvdH score, total JSN score, and total erosion score, each summed over all relevant hand and foot joints of a given patient at a given point in time. B. Confusion matrices of individual joint scores, grouped for JSN and erosion as well as hand and foot joints, respectively. Numbers within the colored fields represent the fraction of joints with a given ground truth score (x axis) and a predicted score (y axis) with respect to the total number of joints with that given ground truth score. Bars to the top and right of each confusion matrix indicated the total number of joints with a given ground truth and predicted score, respectively. ρ, spearman correlation; ICC, intraclass correlation.

**Figure 3.**
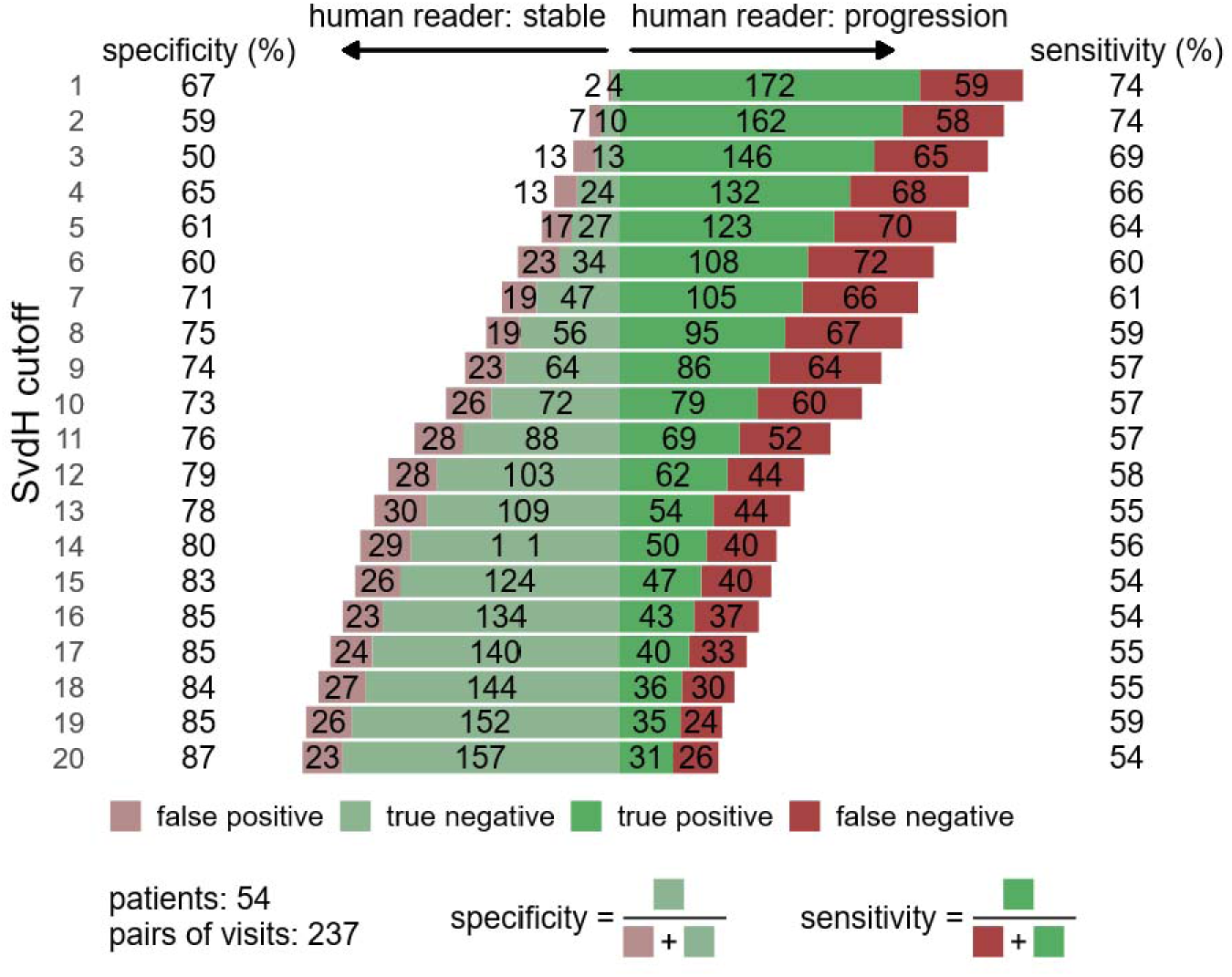
Detection of radiographic progression compared to ground truth human reader for different SvdH cutoffs (y axis). Numbers within the bars represent pairs of visits. Green bars indicate agreement between the automated scoring system and the human reader while red bars indicate disagreement. Bars towards the left of the center (faded colors) indicate no progression (stable disease) according to the human reader while bars to the right indicate progression (full colors). Specificity and sensitivity can be read off the chart by comparing green to red bars towards the left and right, respectively, as indicated in the legend below the bar chart. Precalculated values for specificity and sensitivity for each row are given in the columns to the left and right of the bar chart, respectively.

### Localization and ROI extraction

Filtering for images that showed no operated joints and, for the localization task, had available pixel spacing values, 91 hands and 93 feet of 79 patients were available for localization as well as 94 hands and 95 feet of 80 patients for ROI extraction. Supplementary Figure 5 displays the results of the joint localization network and the ROI extraction algorithm evaluated for deviation of joint locations and presence of overlapping or cropping of extracted ROIs, respectively. ROIs were also examined for malrotation of more than 30° but such artifacts did not occur in a single case and are therefore not shown in Supplementary Figure 5. The results revealed accurate localization of ROIs in most joint groups. For the wrist, an inconsequentially increased deviation is expected due to the larger ROI size. For ROI extraction, the manual quality assessment revealed very few overlap or cropping artifacts. The frequent overlap for CMCs is unavoidable due to how CMCs appear on plain radiographs.

## Discussion

Monitoring for radiographic joint damage is a key component in the management of patients with rheumatoid arthritis and serves as an important outcome measure in clinical trials. However, its semi-quantitative assessment by validated scoring systems is hampered by practical limitations in the form of time and human resources constraints as well as by interrater variability. To circumvent and alleviate those restrictions, we have developed autoscoRA, a fully automated scoring system leveraging a deep learning-based approach to assign SvdH scores to sets of radiographs consisting of both hands and feet.

Early previous attempts generally used very small data sets and were limited by the application of more traditional computational methods, while newer methods such as deep learning tend to rely on large amounts of input data and have only recently begun to be applied. Rohrbach et al. reported on a promising initial deep learning-based approach to automating the Ratingen Rau score^37^ on a very large dataset, but their system only included MCP and PIP joints of the hands, did not feature automatic detection and segmentation of joints, and it has not been validated for its ability to detect progression^38^. A recent machine learning challenge allowed teams of computer scientists globally to develop and test their own approach to automating the SvdH score on mostly cross-sectional data including 674 sets of radiographs with an additional validation cohort consisting of 50 sets of images^39^. While an exciting approach to “crowdsource” the development of an automated scoring system, the (validation) data set used was still relatively small, the ranking and validation of submitted models was not based on intuitive clinically relevant criteria, and given the lack of sufficient longitudinal data, there was no validation for the detection of radiographic progression over time.

The system we present automatically localizes joints, extracts regions of interest around joints for further processing, and finally assigns SvdH scores for both JSN and erosion to each joint. It achieved excellent agreement with the original human reader on the hitherto largest independent test set and outperformed a second human reader on a subset of that data set. While neither the focus of this work, which attempted to emulate the assignment of absolute SvdH scores, nor a straightforward task given that the original human reader performed parallel scoring (i.e., scoring progression by comparing two subsequent radiographic sets) while the automated system only saw individual sets of radiographs at a time, the system also managed to achieve good agreement with the ground truth for radiographic progression. Progression detection also showed stable performance independent of which SvdH cutoff was used to define significant radiographic progression.

A major challenge in training an automated scoring system consists in the inherent imbalance of the underlying scoring data, with little data available for very severely affected joints. We attempted to mitigate this problem by oversampling our training data. Another consequence of the rare occurrence of higher joint-level scores is that some joint-level erosion scores of the feet simply were not present in our test set.

We chose to use an ROI containing the entire wrist for each model predicting a wrist joint score. Each wrist joint score was still predicted individually to avoid losing granularity. The advantage of this approach is that we avoided the potentially error-prone process of cutting out specific parts of the wrist, making the pipeline more robust. In addition, due to wrist anatomy as visible on radiographs, such subregions of the wrist would necessarily still overlap to a significant degree, limiting the advantage of extracting them individually. However, it meant that each model needed to learn from the data which part of the wrist was of interest, arguably making the task harder.

Another limitation pertains to the establishment of ground truth labels. While consensus scores from two radiographers are often used to approximate ground truth, the present study relied on labels from a single expert scorer for training. To mitigate this, a second scorer—trained by the primary expert—was included for evaluation, thereby providing an additional independent assessment. This also enables interpreting the difference between the model and reader 1 vs. the model and reader 2 in light of reader 1 providing values for algorithm training. Despite this limitation, the proposed framework offers a scalable approach for the automated scoring of RA X-rays, trained and validated using annotations from a single scorer, and demonstrated superior performance compared to a second, similarly trained human scorer. The fact that reader 2 was trained by reader 1 could lead to a low estimate of inter-reader variability, making the models’ outperformance of reader 1 in approximating reader 2 scores a strong argument for the increased consistency provided by an automated scoring system.

A limitation of this study is the lack of external validation on data from independent institutions. Future work will include multi-center external datasets to further assess model generalizability and clinical applicability.

A further limitation is that, although the model outputs joint-level scores and thus highlights the specific regions driving patient-level predictions, it does not yet provide pixel-level interpretability. Saliency maps could enhance this aspect by visualizing the precise image features contributing to each joint score.

Limiting the application of our automated radiographic scoring system in routine clinical care, where even small changes might be of relevance, is that it was designed primarily to quantify absolute structural damage rather than natively comparing two consecutive radiographs (as a radiologist would).

In summary, the clinical utility of autoscoRA extends across the research–practice continuum. In clinical trials, automated SvdH scoring can reduce costs, shorten timelines, and eliminate reader variability, thereby enhancing the precision and interpretability of radiographic endpoints. In registries and observational studies, it enables large-scale transformation of raw radiographs into structured quantitative data, facilitating robust analyses of long-term outcomes. Looking forward, in routine rheumatology practice, where systematic scoring has been largely unfeasible, such automation opens the possibility of integrating structural progression as a practical and standardized component of disease monitoring. While external validation and direct progression-focused training remain important next steps, autoscoRA represents a pivotal advance toward making radiographic scoring not only more accurate and reproducible but also truly accessible in everyday clinical and research settings.

## Data Availability Statement

The data utilized in this study consist of retrospective patient data. Due to privacy regulations and institutional policies, these data cannot be made publicly available in a global repository. However, access to the data may be granted upon reasonable request and subject to a data transfer agreement (DTA) with the respective institutions.

## Acknowledgement

The authors would like to acknowledge the help of Gabriella Supp in identifying imaging data sets and manual scoring of radiographs.

## Footnotes

a Model implementation is available at: https://github.com/weiserjpaul/AutoScorRA

