## Supplements for "autoscoRA: Deep Learning to Automate Sharp/van der Heijde Scoring of Radiographic Damage in Rheumatoid Arthritis"

Authors

Thomas Deimel^1,^*, Paul J. Weiser^2,^*, Martin Urschler^3^, Christian Payer^4^, Georg Langs^2^, Daniel Aletaha^1^.

1.) Division of Rheumatology, Department of Medicine, Medical University of Vienna, Vienna, Austria.
2.) Computational Imaging Research Lab, Department of Biomedical Imaging and Image-guided Therapy, Medical University of Vienna, Vienna, Austria.
3.) Institute for Medical Informatics, Statistics and Documentation, Medical University of Graz, Graz, Austria
4.) Institute of Computer Graphics and Vision, Graz University of Technology, Graz, Austria

*.) Authors contributed equally

Supplement

Distribution of human reader SvdH scores

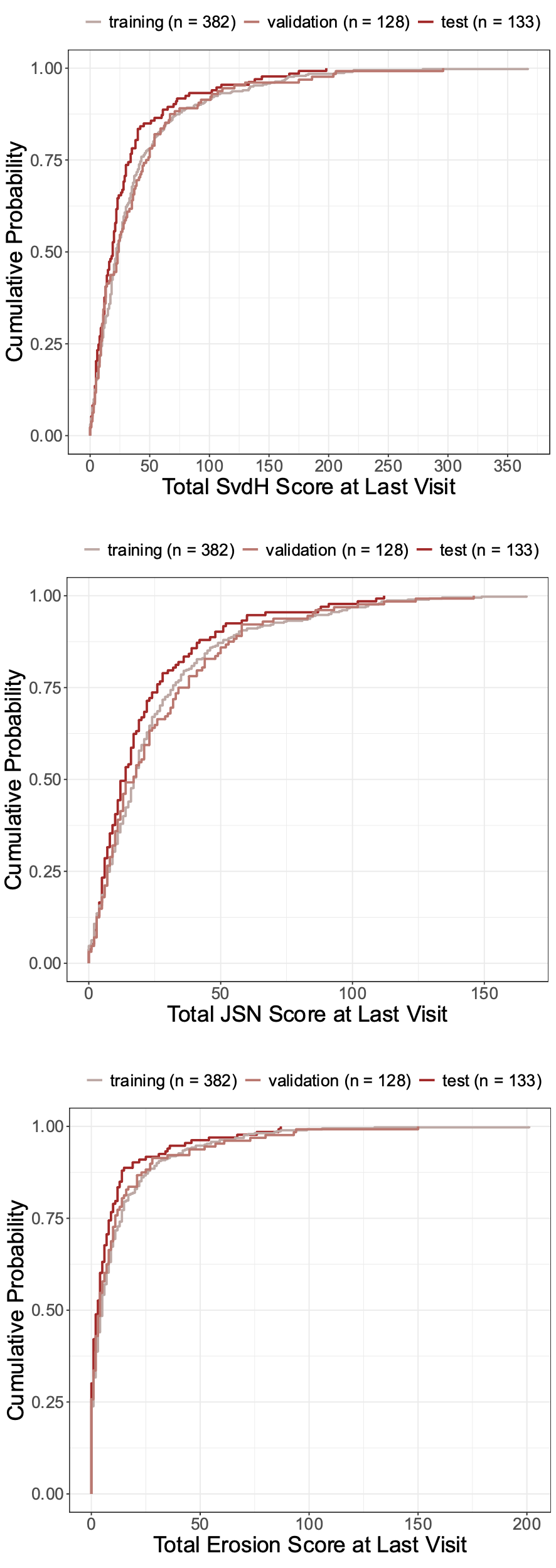

Supplementary Figure 1. ECDF for SvdH, Erosion, and JSN score assigned by the primary human reader at patients’ last visit with a complete set of radiographs (two hands and two feet), stratified by dataset. ECDF, empirical cumulative distribution function.

Automated scoring of total SvdH across different levels of joint damage

Supplementary Table 1. Performance of automated scoring of total SvdH scores across different levels of human reader-assessed joint damage. MAE, mean absolute error; RMSE, root mean square error.

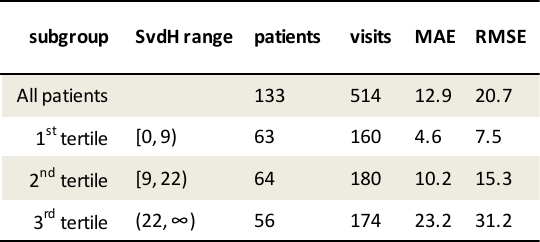

Human vs. machine learning

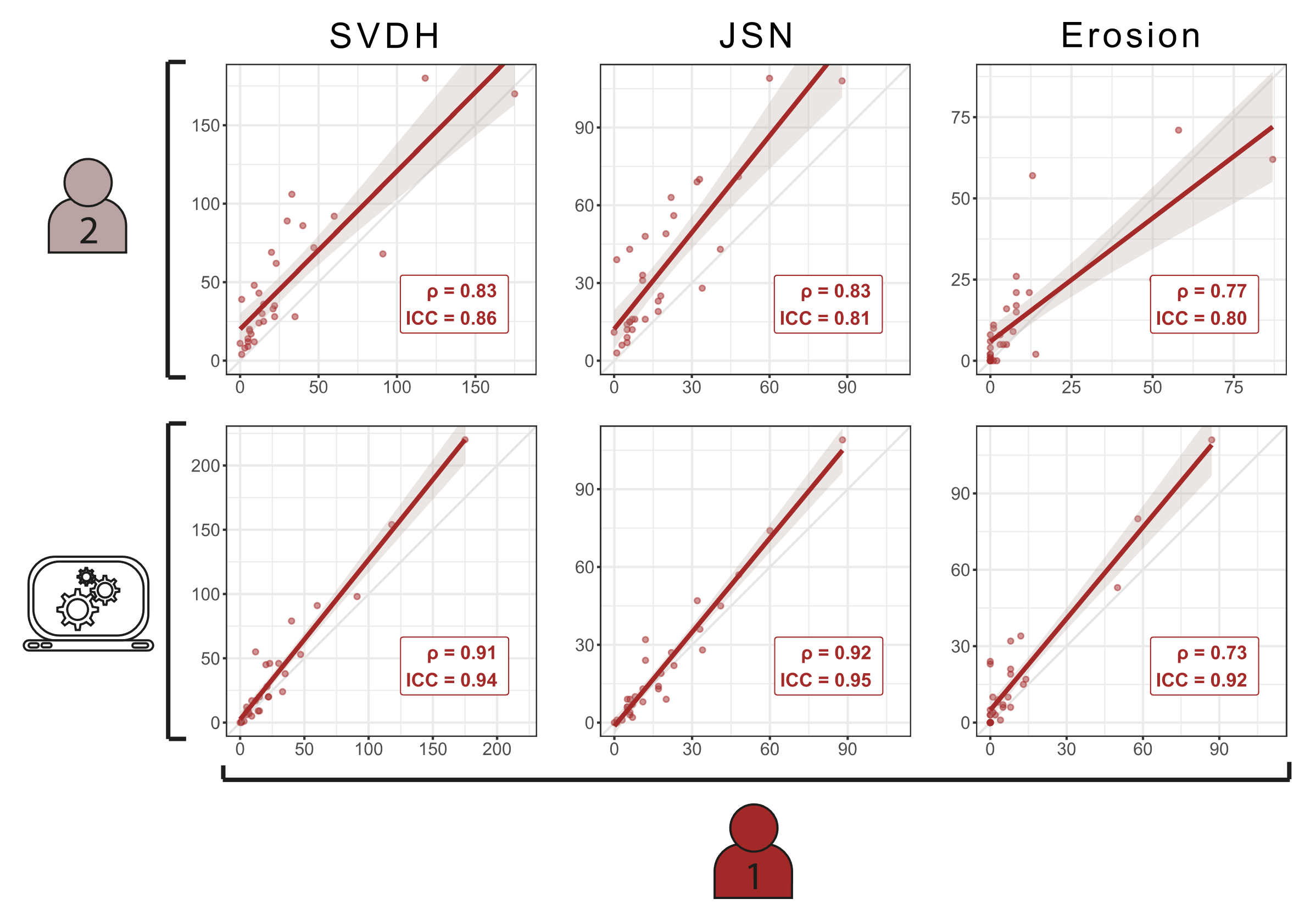

Supplementary Figure 2. Comparison between the automated scoring system and a second human reader. Scatter plot of sum scores, including the total SvdH score, total JSN score, and total erosion score, each summed over all relevant hand and foot joints of a given patient at a given point in time. ρ, spearman correlation; ICC, intraclass correlation.

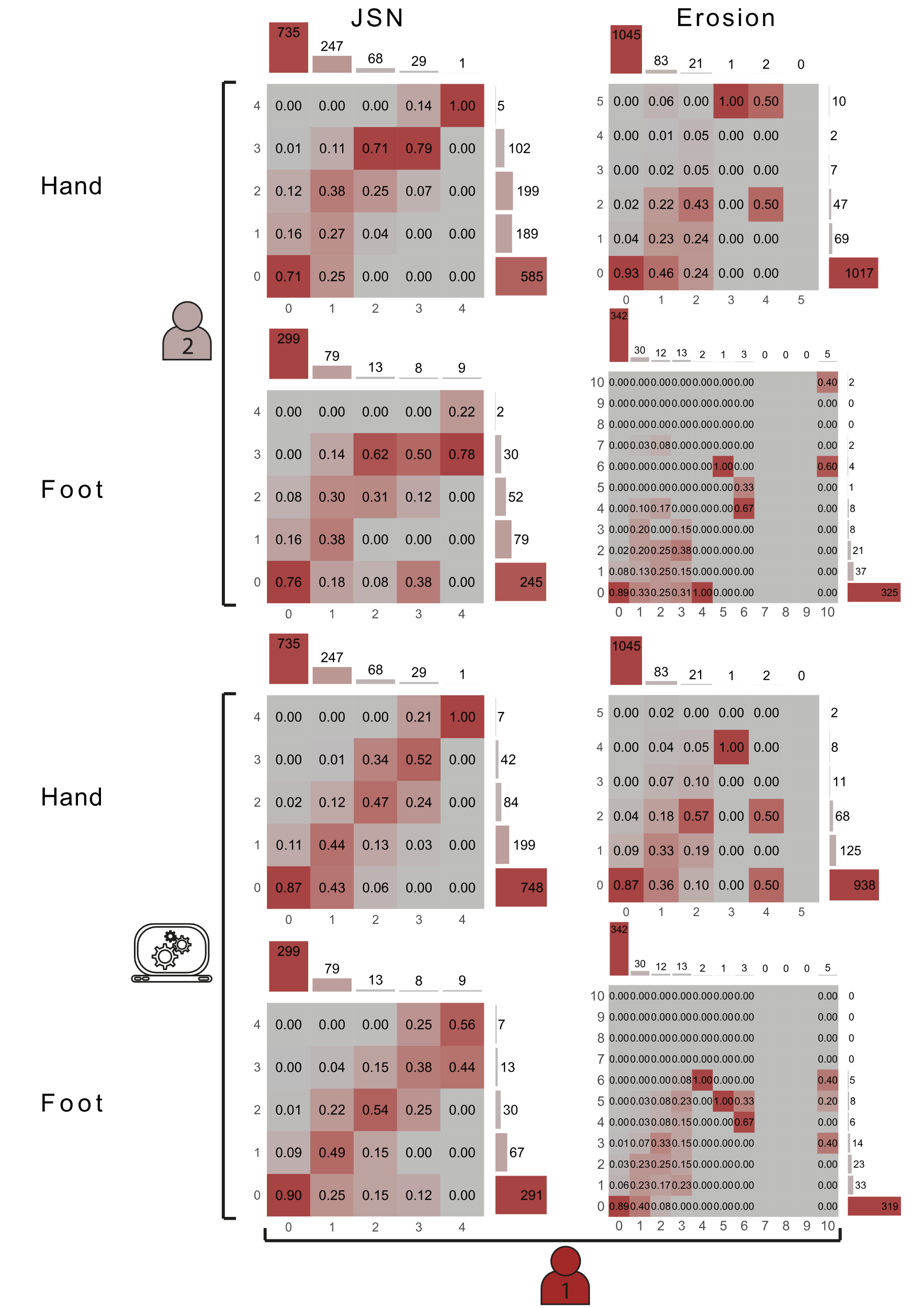

Supplementary Figure 3. Comparison between the automated scoring system and a second human reader. Confusion matrices of individual joint scores, grouped for JSN and erosion as well as hand and foot joints, respectively. Numbers within the colored fields represent the fraction of joints with a given ground truth score (x axis) and a predicted score (y axis) with respect to the total number of joints with that given ground truth score. Bars to the top and right of each confusion matrix indicated the total number of joints with a given ground truth and predicted score, respectively.

Radiographic progression at different degrees of baseline joint damage

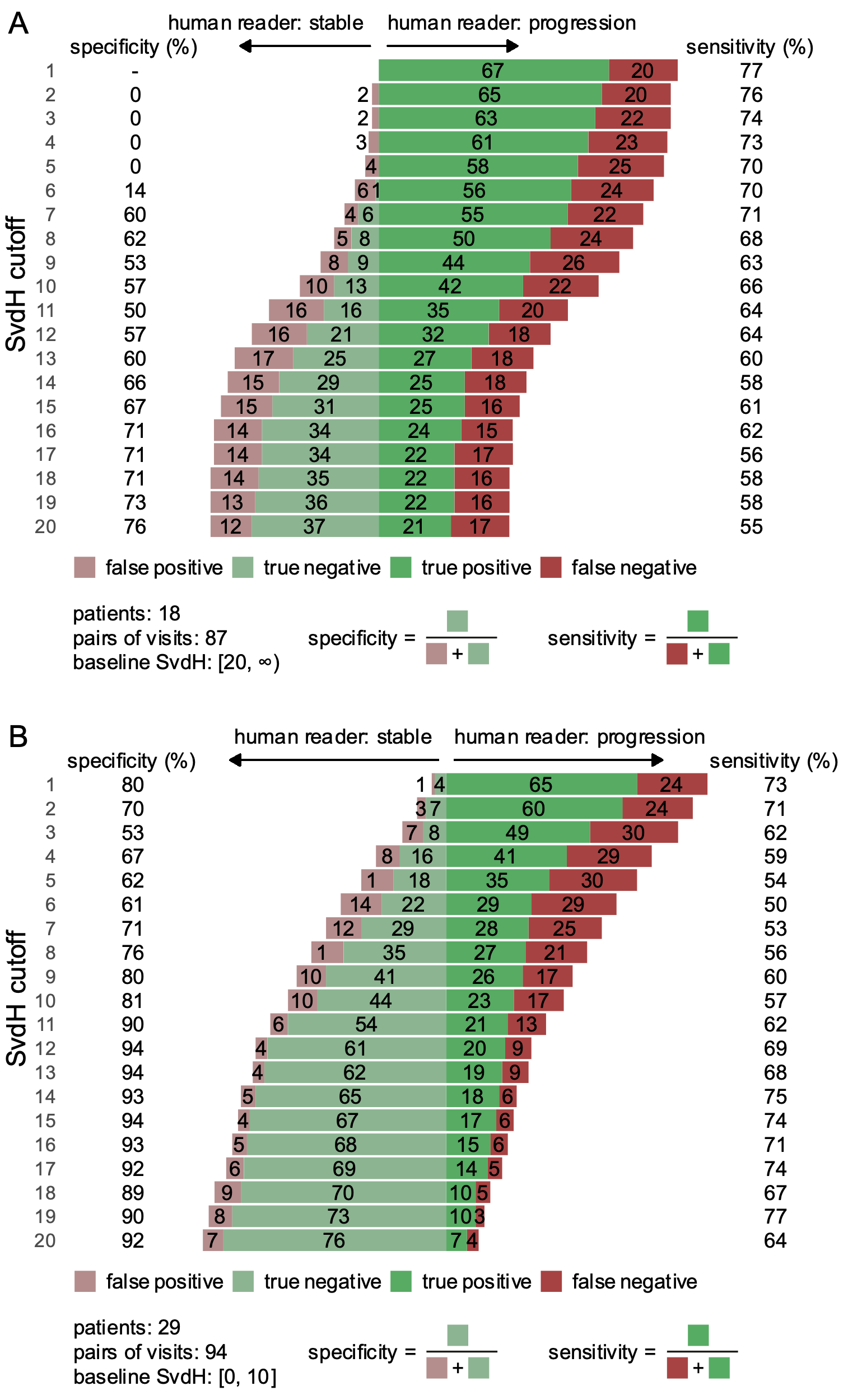

Supplementary Figure 4. Detection of radiographic progression compared to ground truth human reader for different SvdH cutoffs (y axis). A. Baseline human reader SvdH score ≥ 20. B. Baseline human reader SvdH score ≤ 10.

Localization and ROI extraction

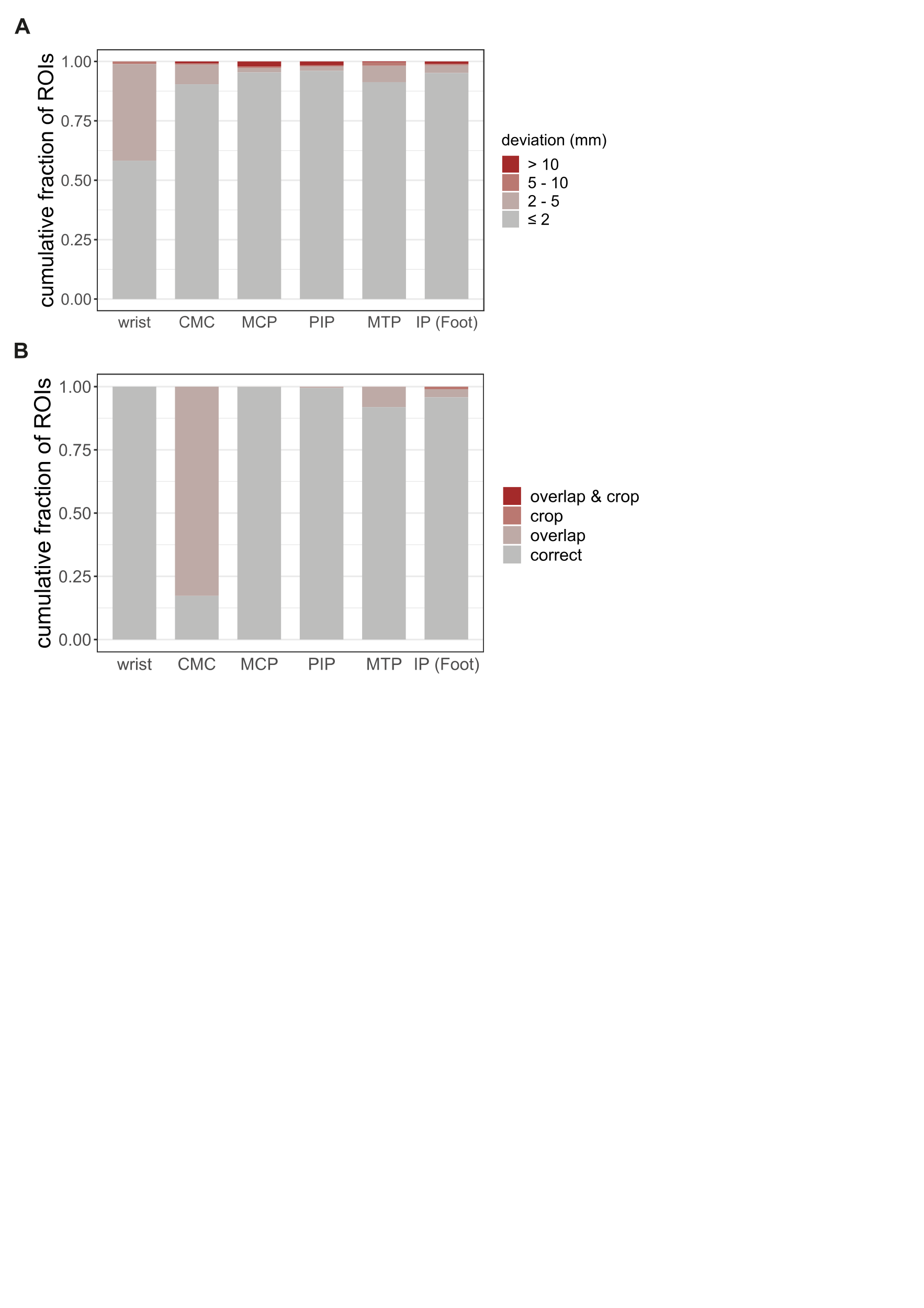

Supplementary Figure 5. A. Localization. Three-fold cross validation results for automated joint localization using the SpatialConfiguration-Net^32^. The frequent occurrence of small deviations during localization of the center of the wrist region of interest is unlikely to have affected further steps since the size of the region of interest easily compensates for slight variations in its position. B. ROI extraction. Three-fold cross validation results for ROI extraction based on whether the entirety of the joint space and bone of interest was depicted (no “crop”) and on whether parts of any adjacent joint spaces, other than that of the joint of interest, were visible (“overlap”). The frequent overlap for CMCs is unavoidable due to how CMCs appear on plain radiographs. CMC, carpometacarpal joints; MCP, metacarpophalangeal joints; PIP, proximal interphalangeal joints (including IP joint of the thumb); MTP, metatarsophalangeal joints; IP, interphalangeal joint of the first toe.
